# COVID-PCD – a participatory research study on the impact of COVID-19 in people with Primary Ciliary Dyskinesia

**DOI:** 10.1101/2020.11.11.20229922

**Authors:** Eva SL Pedersen, Eugénie NR Collaud, Rebeca Mozun, Cristina Ardura-Garcia, Yin Ting Lam, Amanda Harris, Jane S Lucas, Fiona Copeland, Michele Manion, Bernhard Rindlishbacher, Hansruedi Silberschmidt, COVID-PCD patient advisory group, Myrofora Goutaki, Claudia E Kuehni

## Abstract

**Introduction:** COVID-PCD is a participatory study initiated by people with PCD who have an essential vote in all stages of the research from the design of the study to the recruitment of participants, and interpretation and communication of the study results. COVID-PCD aims to collect epidemiological data in real time from people with PCD throughout the pandemic to describe incidence of COVID-19, symptoms, and course of disease; identify risk factors for prognosis; and assess experiences, wishes, and needs.

**Methods:** The study is advertised through patient support groups and participants register online on the study website (www.covid19pcd.ispm.ch). The study invites persons of any age from anywhere in the world with a suspected or confirmed PCD. A baseline questionnaire assesses details on PCD diagnosis, habitual symptoms, and COVID-19 episodes that occurred before study entry. Afterwards, participants receive a weekly follow-up questionnaire with questions on incident SARS-CoV-2 infections, current symptoms, social contact behaviour, and physical activity. Occasional thematic questionnaires are sent out focusing on emerging questions of interest chosen by people with PCD. In case of hospitalisation, patients or family members are asked to obtain a hospital report. Results are continuously analysed and summaries put online.

**Conclusion:** The study started recruitment on April 30, 2020, and 556 people with PCD completed the baseline questionnaire by November 2, 2020. The COVID-PCD study is a participatory study that follows people with PCD during the COVID-19 pandemic, helps to empower affected persons, and serves as a platform for communication between patients, physicians, and researchers.

## Introduction

The pandemic of coronavirus disease 2019 (COVID-19), caused by SARS-CoV-2 has by November 2020 spread globally with more than 47 million infected people in over 200 countries and territories. The severity of COVID-19 ranges from asymptomatic to severe disease (1, 2), and elderly people and individuals with pre-existing chronic health conditions are at increased risk of a severe disease course (3-7). COVID-19 has also been associated with impaired lung function after recovery, even in people without pre-existing health conditions (8, 9).

Primary Ciliary Dyskinesia (PCD) is a rare genetic multi-system organ disease that affects about 1 in 10.000 people. It leads to chronic upper and lower airway disease, laterality defects including congenital heart disease, and other health problems (10-13). While some patients are relatively mildly affected, others develop severe irreversible lung disease from an early age with repeated infections resulting in reduced lung function, bronchiectasis, and sometimes oxygen dependency with some requiring lung transplantation (14-19). We do not know how SARS-CoV-2 affects people with PCD and whether severity of PCD, age, and other patient characteristics influence COVID-19 disease course. PCD registries and research databases are not designed for a longitudinal collection of patient-reported information on symptoms and health behaviour in real time (20). However, such data are needed to comprehensively understand the impact of SARS-CoV-2 on people with PCD. Patient-reported data are also essential to understand how public health measures that aim to reduce the spread of COVID-19 are adopted by people with PCD, and how the pandemic affects their life and medical care. During the early phase of the pandemic (March and April, 2020), people with PCD all over the world were worried and desperate to understand effects of SARS-CoV-2 on persons with this rare disease. Given the lack of evidence, patient support groups approached researchers and proposed to set up a study together.

Research projects that focus on rare diseases can be challenging due to the limited number of people with the disease and their geographical dispersion (21). It is therefore important to make the study as relevant as possible to the participants and ensure that participation is feasible. This can be helped by using a participatory approach in which study participants are engaged in all stages of a research project from the design of the study to shaping the content and interpreting results. Participatory research methods have been associated with better networking between participants, higher relevance for affected people, patient empowerment, and higher participation rates (22-24). COVID-PCD has been designed as a collaboration of epidemiologists, health care specialists, and people with PCD. It is a longitudinal study aiming to collect data in real time on health, health care, behaviour, and psychosocial aspects from people with PCD throughout the COVID-19 pandemic. This manuscript describes aims, methods, and first results of COVID-PCD.

## Methods

### Study objectives

We set up an online surveillance system which collects essential epidemiological and clinical data in real-time directly from people with PCD and publishes continuously summarized results. The study aims to:

1. Describe incidence of COVID-19 in people with PCD, symptoms, course of disease, duration of illness, treatments, and outcomes;
2. Identify risk factors for incidence and prognosis;
3. Describe the impact of the pandemic on daily life and health care in people with PCD, and;
4. Assess patients’ experiences, wishes, and needs during the COVID-19 pandemic.

### Study design

COVID-PCD is an international observational cohort study using anonymous online questionnaires to collect information directly from people with PCD (clinicaltrials.gov registration number: NCT04602481). It is a participatory research project where people who have PCD have an active role in all stages of research. The study was requested by PCD support groups from Switzerland, Germany, the United Kingdom, the USA, and Australia and set up in collaboration with a study team at the University of Bern, Switzerland, and the University of Southampton, UK. The study is designed for three age groups; children below 14 years, adolescents between 14 and 17 years, and adults aged 18 years or more. For children, the questionnaires are addressed to the parents, but the child is encouraged to help complete the questionnaires. Adolescents complete the questionnaires themselves. The study is currently available in English, German, and Spanish with the possibility to expand to more languages.

### Inclusion criteria

The study is addressed to people of any age from any country with a confirmed or suspected diagnosis of PCD. Eligibility of participants is assessed in the baseline questionnaire. We exclude people who answer ‘no’ to both questions: ‘Have you been told by a doctor that you have PCD or are likely to have PCD?’ and ‘Do you have a reason to believe that you have PCD, even though you have not received a diagnosis by a doctor?’.

### Validating the diagnosis of PCD

Diagnosing PCD is complex and requires several tests to be performed by an experienced team. Historically, patients may not have been diagnosed according to current standards (25) and in some patients, even experts are unable to confirm the diagnosis. We validate the diagnosis through questions asking the patients about results of the diagnostic tests. Many patients have received copies of their test results and can therefore report the type of tests they have had and their results. For the analysis, we can stratify the study population into people with a definite diagnosis, people with probable PCD, and people with suspected PCD. During the study, we will further attempt to validate the diagnoses with original documents. We considered it important to have an inclusive approach and allow also people whose diagnosis is not proven into the study, because whether or not participants have a formal confirmation of PCD, they suffer from chronic lung disease, and thus should be offered to contribute their experiences.

### Recruitment of study participants

The study is primarily advertised through PCD support groups (**table 1**) who contact and inform potential participating people with PCD through social media and email networks and encourage them to take part in the study. The adverts include a link to the project website (www.covid19pcd.ispm.ch) which leads directly to the study information and registration. Initially, support groups from the UK (www.pcdsupport.org.uk), Germany (www.kartagener-syndrom.org/index.php), Switzerland, the USA and Canada (www.pcdfoundation.org), and Australia (www.pcdaustralia.org.au/) were involved, but more countries join as the study progresses. In regions of the world where PCD specific patient support groups are lacking, health care specialists seeing people with PCD are asked to help inform patients about the study.

**Table 1:** Collaborating PCD support organisations

| PCD support organisation | Country | Contact persons | Website |
| --- | --- | --- | --- |
| **Selbsthilfegruppe Primäre Ciliäre Dyskinesie** | Switzerland | Bernhard Rindlisbacher | - |
| **Verein Kartagener-Syndrom und PCD** | Germany | Hansruedi Silberschmidt | www.kartagener-syndrom.org/index.php/verein |
| **PCD Family Support Group UK** | United Kingdom | Lucy Dixon | www.pcdsupport.org.uk |
| **PCD Foundation** | USA and Canada | Michele Manion | www.pcdfoundation.org |
| **PCD Australia Primary Ciliary Dyskinesia** | Australia | Catherine Kruljac | www.pcdaustralia.org.au/ |
| **Associazione italiana Discinesia Ciliare Primaria Sindrome di Kartagener Onlus** | Italy | Letizia Andolfi, Sara Bellu | www.pcdkartagener.it/associazione/ |

### Study procedures

All aspects of the COVID-PCD study are online, including registration and consent and completing the baseline questionnaire, weekly questionnaires, information on hospitalisation, and occasional thematic questionnaires (**figure 1**). The website (www.covid19pcd.ispm.ch) includes detailed information about the study results and allows participants to register via a link that leads them directly into the study database. At first, participants are asked to enter their year of birth which directs them to the age-specific study information and questionnaires. Participants give digital consent to participate and are then asked to enter an email address where the online questionnaires can be sent to. Immediately after the registration, participants receive an email with the link to the baseline questionnaire. Adolescents are asked to also enter the email address of a parent. The parent must give consent before the adolescent receives the baseline questionnaire. Participants must complete the baseline questionnaire to be enrolled in the study.

**Figure 1:**
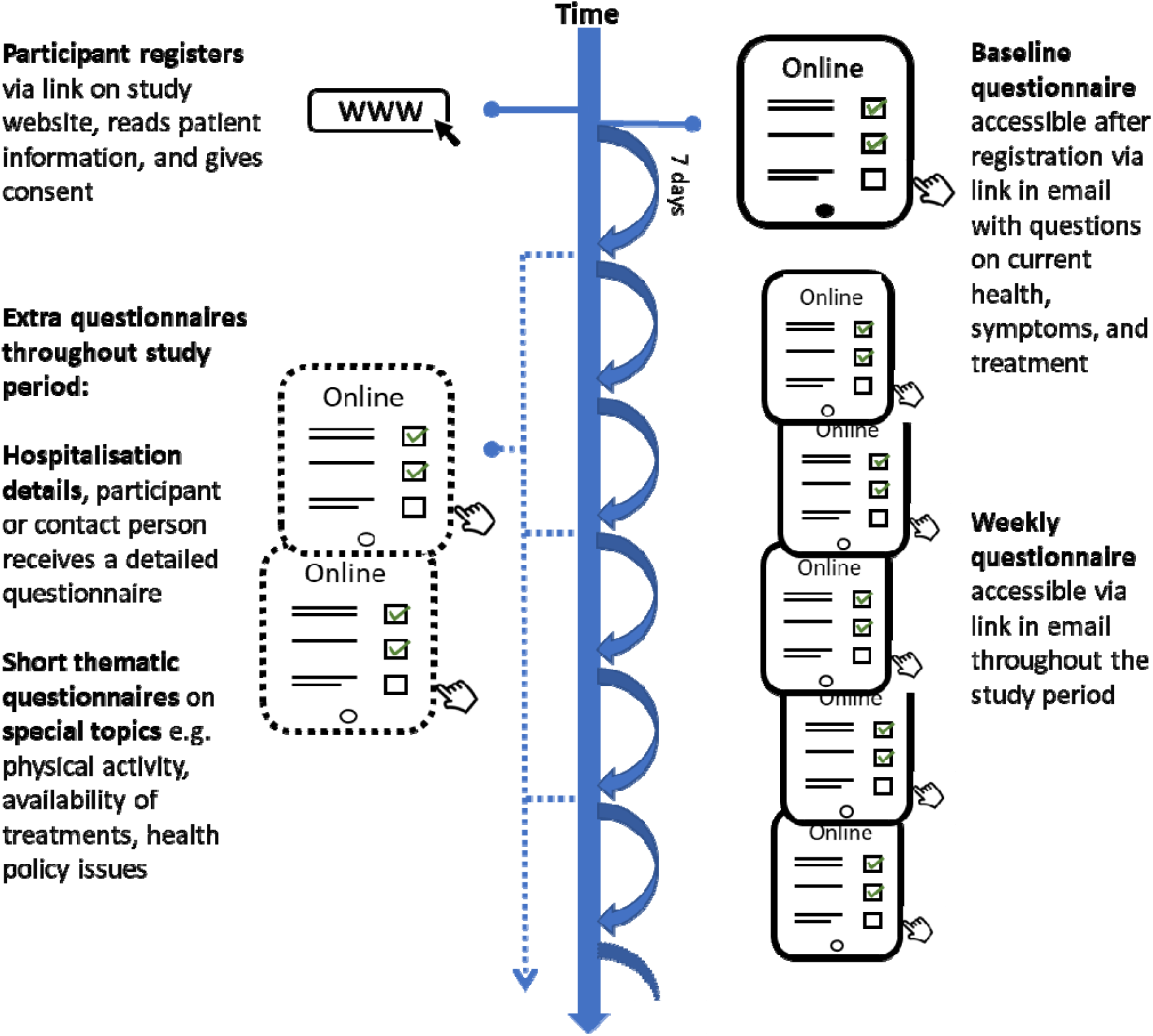
Timeline and follow-up procedures of the COVID-19 and PCD study.

One week after completing the baseline questionnaire, and in weekly intervals thereafter, participants receive short follow-up questionnaires (**figure 1**). The follow-up questionnaire is not dependent on participants completing it every week; if participants misses one or more of the follow-up questionnaires, they will receive the links for the next ones. If a participant is hospitalised due to COVID-19, a link to an additional short questionnaire is sent. The study allows also to conceive and send out thematic questionnaires that focus on specific topics related to the COVID-19 pandemic. Participants and interested people can follow the progress of the study on the study website where information on number of recruited participants, incident COVID-19 infections, and selected results from the questionnaires are uploaded regularly.

### Information collected

**The baseline questionnaire** aims to assess whether the participant has PCD, how the disease was diagnosed, what the person’s usual symptoms are, how severe the disease is, and how it is treated. It also aims to collect information on the social and environmental context such as people living in the same household, school and work environment, and physical activity. The baseline questionnaire is divided into four modules (**table 2**). Module 1 collects demographic data, contact information, and information on SARS-CoV-2 infections that have occurred prior to study entry. Module 2 collects information on PCD diagnosis and baseline disease status. Module 3 asks about regular PCD-related symptoms, and module 4 collects information on work, education, lifestyle, and health behaviours. Module 3 and 4 are based on the FOLLOW-PCD questionnaire (26).

**Table 2:** Information collected in the COVID-19 and PCD study

| Questionnaire | Timing of distribution | Information collected |
| --- | --- | --- |
| **Registration questionnaire** | At registration | Language, year of birth, consent to participate, Email address |
| **Baseline questionnaire** | Right after registration | **Module 1:** Demographic information (country, region); contact information in case of hospital stay; history of COVID-19 infection (tests, test results, severity of infection, symptoms, people in household who had a COVID-19 infection), other infections.<br>**Module 2:** PCD diagnosed by a physician (study eligibility question), PCD in close family, prior diagnostic tests for PCD and test results (nasal nitric oxide, high speed video microscopy, electron microscopy, genetic test), year of PCD diagnosis, situs inversus, congenital heart defect, bronchiectasis, infections (bacteria, viruses, fungi), lung function, height, weight, airway clearance therapy, physiotherapy, oxygen use, antibiotic treatments, hospital stays, flu vaccinations. Comorbidities and their treatments including asthma, hypertension, diabetes, heart failure, primary immunodeficiency, cancer, inflammatory bowel disease, and stroke.<br>**Module 3:** (adapted from the standardized FOLLOW-PCD patient questionnaire*): Current symptoms: Symptoms from nose or ears, hearing problems, headaches, snoring, coughing, coughing up mucus, wheezing, shortness of breath, chest pain, fever, general feeling of being unwell.<br>**Module 4:** (adapted from the standardized FOLLOW-PCD patient questionnaire*): Employment/educational status, profession, days missed from work due to PCD, mode of transportation to work/school, highest level of education. Level of physical activity, types of sports/activities regularly performed, smoking status, number of |
|  |  | cigarettes per day, years of smoking, year of quitting smoking, anyone else in household smoking, nutrition (appetite in last 3 months, additional drink supplements, other supplements), living conditions (Type of home, number of bedrooms in home, access to garden/yard), change in living condition due to COVID-19 pandemic. |
| **Follow-up questionnaire** | Weekly (period can be adapted) | Current symptoms (in the last 7 days).<br>Recent COVID-19 infections: Contact with health professional due to COVID-19 infection, medication taken in the past 7 days, change in usual therapies past 7 days.<br>Contact behaviour in the past 7 days: Activities (e.g. stayed at home, went grocery shopping, went out for exercise, went occasionally or regularly to workplace/school, etc.), use of public transportation, possible to keep 2 metres distance to other people during public transport/at work/at school, number of people having close contact with. |
| **Hospitalisation questionnaire** | If a participant is hospitalised | Date of hospitalisation, hospital unit in which patient was treated, duration of hospital stays, treatments, recommendation after discharge. |
| **Thematic questionnaires** | In irregular intervals as research questions arise | Potential topics:<br>Availability of PCD treatments during COVID-19 pandemic<br>Physical activity during COVID-19 pandemic<br>Protection measures at schools and workplaces e.g. masks, plastic face shields, gloves, etc.<br>Anxieties and fears related to the COVID-19 pandemic |

\*Goutaki M, Papon JF, Boon M, et al. Standardised clinical data from patients with primary ciliary dyskinesia: FOLLOW-PCD. *ERJ Open Res.* 2020;6(1).

**The weekly follow-up questionnaire** collects information on symptoms and incident SARS-CoV-2 infections, physical activity, and time-varying lifestyle factors such as social contacts. If a participant has no symptoms, the weekly follow-up questionnaire is short. If a participants has symptoms, extra questions appear asking details about tests for SARS-CoV-2 and other infections, symptoms duration, health care contacts, and medication taken.

**A short questionnaire is sent in the event of a hospitalisation** due to COVID-19. It asks about duration of the hospitalisation, unit where the patient was treated, treatments, and recommendations received after discharge.

**Additional thematic questionnaires** developed throughout the study focus on special topics related to the pandemic such as mask use, anxiety, home schooling of children, physical activity, or availability of medications. PCD support organisations and study participants are encouraged to suggest special topics and participate in developing those questionnaires.

### Participant engagement

Participants shape the study in a participatory approach and can influence all aspects of the study (**table 3**). They had a decisive role in study design and content of baseline questionnaire. As the study goes on, participants are encouraged to propose research questions and topics for extra questionnaires, help to make sure questions are written in a language that is easily understandable, advise on logistics such as how to approach affected people, and choose results for publication on the website. A patient advisory group from the European Lung Foundation (ELF, www.europeanlung.org/en/), includes interested people who have PCD. The group meets to discuss the COVID-19 outbreak, the PCD-COVID study, and questions relevant to people with PCD.

**Table 3:** Participant involvement in the study

| Study step | Inputs from PCD support groups |
| --- | --- |
| Concept and design of study | • The idea for the study came from PCD support groups<br>• The study design was developed in close communication between PCD support groups and the study team |
| Questionnaire design | • The content of the baseline and follow-up questionnaires was decided based on ideas from PCD support group representatives, health care staff and researchers<br>• Wording of the questionnaires were checked by PCD support group representatives to ensure that the language was understandable and meaningful for all |
| Recruitment of participants | Study participants are recruited using measures such as:<br>• PCD support groups social media post links to the study website<br>• Emails to PCD support group members<br>• Presentations of the study at PCD support group meetings and in newsletters |
| Choice and interpretation of results | Summarised results displayed on the website are based on what is most important for participants |
| Thematic questionnaires | Thematic questionnaires are developed based on suggestions from participants and PCD support groups |
| Display of results on study website | Participants can suggest results to be posted on the website and how to phrase them |

### Study database and data protection

The web-based database uses the Research Electronic Data Capture (REDCap) platform developed at Vanderbilt University (www.project-redcap.org) (27). The database is hosted by the Swiss medical registries and data linkage centre (SwissRDL) at the University of Bern, Switzerland, and complies with all legal requirements for data security and data protection. Access to the database is regulated by a system with personal passwords. Only study team members who need to perform database management can access the database. Daily, weekly, and monthly back-ups are performed and securely stored at the SwissRDL servers.

### Ethics

The Bern Cantonal Ethics Committee (Kantonale Ethikkomission Bern) has approved this study (Study ID: 2020-00830). Informed consent to participate in the study is provided at registration into the study. Study participation is anonymous. Participants can withdraw their consent to participate at any time by contacting the study team.

### Current recruitment status

The study started recruitment on April 30^th^, 2020. By November 2, 2020, 556 people with PCD completed the baseline questionnaire (**figure 2**). Together, participants have already completed a total of 4776 weekly questionnaires over a period of 23 weeks. On average, 50% (range 33-68%) of participants completed the weekly follow-up questionnaire (**figure 2**). The study includes people with PCD from 36 countries with the largest numbers from England (n=121; 22%), the USA (n=101; 18%), Germany (n=78; 14%), and Switzerland (n=42, 8%) (**table 4**). Two thirds of participants are female (n=340; 61%) and all age-groups are well represented with 172 (31%) less than 15 years, 136 (24%) aged 15 to 29 years, 154 (28%) between 30 and 49 years, and 94 (17%) being 50 years or older.

**Table 4:** Country of residence and sex of participants, displayed in total and by age groups (N=556)

|  | **Total** | **0-14 y** | **15-29 y** | **30-49 y** | **50 y and above** |
| --- | --- | --- | --- | --- | --- |
|  | N=556 | N=172 | N=136 | N=154 | N=94 |
|  | n(%) | n(%) | n(%) | n(%) | n(%) |
| **Country\*** |  |  |  |  |  |
| England | 121 (22) | 34 (20) | 32 (24) | 36 (23) | 19 (20) |
| USA | 101 (18) | 35 (20) | 23 (17) | 26 (17) | 17 (18) |
| Germany | 78 (14) | 34 (20) | 9 (7) | 21 (14) | 14 (15) |
| Switzerland | 42 (8) | 8 (5) | 16 (12) | 5 (3) | 13 (14) |
| Australia | 27 (5) | 11 (6) | 5 (4) | 5 (3) | 6 (6) |
| Italy | 22 (4) | 7 (4) | 6 (4) | 9 (6) | 0 |
| Canada | 20 (4) | 7 (4) | 3 (2) | 6 (4) | 4 (2) |
| The Netherlands | 18 (3) | 2 (1) | 8 (6) | 5 (3) | 3 (3) |
| Scotland | 12 (2) | 4 (2) | 3 (2) | 3 (2) | 2 (2) |
| Denmark | 10 (2) | 1 (1) | 4 (3) | 2 (1) | 3 (3) |
| France | 10 (2) | 3 (2) | 2 (1) | 2 (1) | 3 (3) |
| Norway | 10 (2) | 2 (1) | 0 | 6 (4) | 2 (2) |
| Other European countries | 60 (11) | 14 (8) | 19 (14) | 22 (14) | 5 (5) |
| Other non-European countries | 25 (4) | 10 (6) | 6 (4) | 6 (4) | 3 (3) |
| **Sex (n=555)** |  |  |  |  |  |
| Male | 214 (39) | 92 (53) | 49 (36) | 41 (27) | 32 (34) |
| Female | 340 (61) | 80 (47) | 86 (63) | 113 (73) | 61 (66) |
| Other | 1 (0) | 0 | 1 (1) | 0 | 0 |

\*Countries with N ≥ 10 displayed in table, countries with N<10 were categorised into other European countries and other non-European countries.

**Figure 2:**
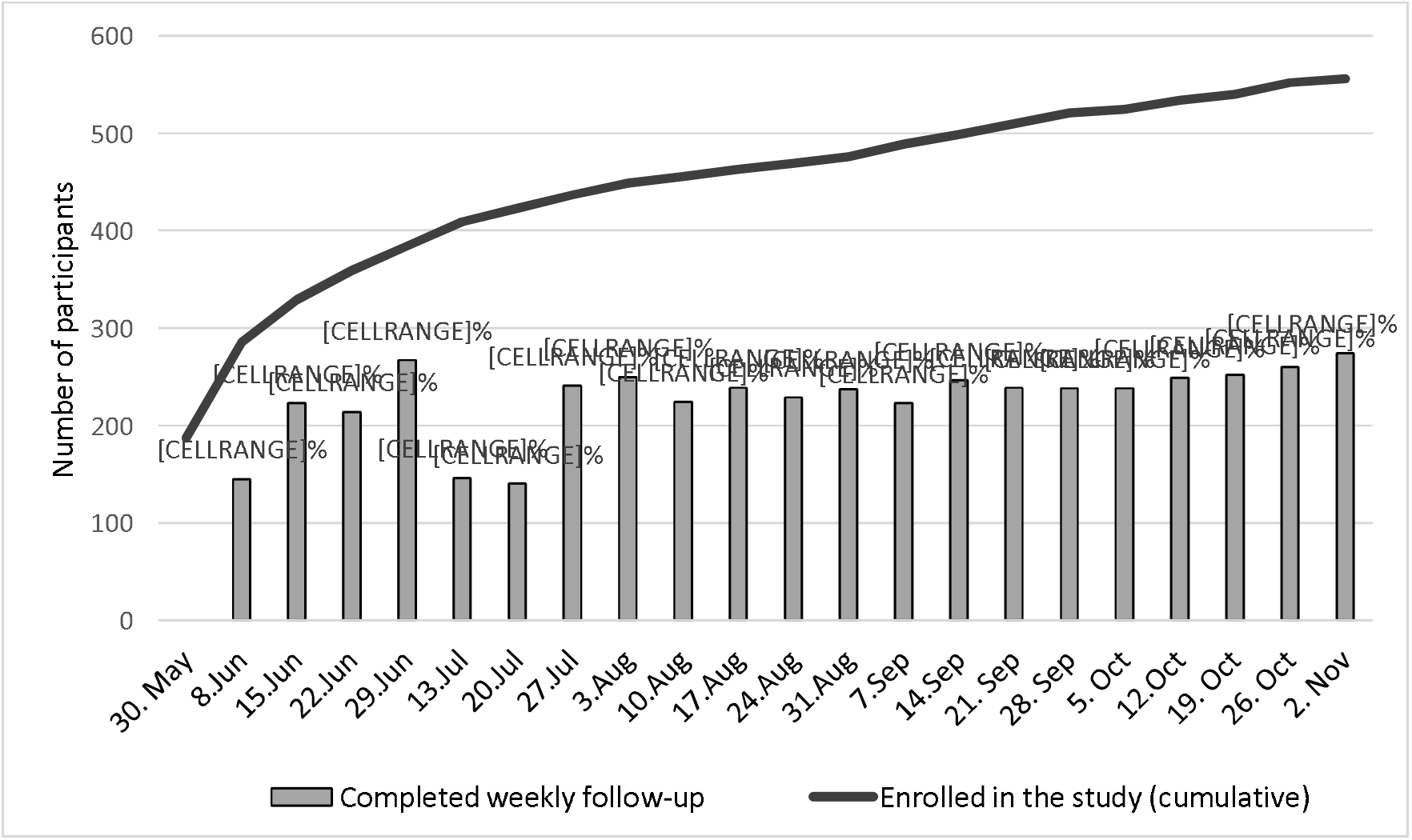
Number of people with PCD who enrolled in the study by November 2, 2020 (cumulative) and number and percent who completed the weekly follow-up questionnaire.

## Discussion

The COVID-PCD study is an online surveillance system of COVID-19 in people with PCD which collects essential information in real-time directly from people with PCD. It is a participatory research project where people who have PCD have an active role in all stages of research. Summarised results are published continuously on the website and shared on social media via the PCD support groups to inform participants, researchers, and physicians. The key features of the study are shown in **text box 1**.

### Text box 1: Key features of the COVID-PCD study

- Participatory study on the effects of COVID-19 in people with PCD
- People with PCD participated to shape the study, design the questionnaires, the recruitment of participants and results to be displayed on the website
- Online cohort study using anonymous questionnaires accessible via link in email
- Inclusion of participants with confirmed or suspected PCD from all over the world
- Weekly online questionnaires capture short-term changes in symptoms, health behaviour and social contact behaviour
- Thematic questionnaires developed throughout the study can focus on specific topics such as mask use, physical activity, and health information policy
- Selected results are published weekly on the study website (www.covid19pcd.ispm.ch)

### Comparison with other studies

To our knowledge, very few studies investigate the effect of COVID-19 on people with a rare disease and very few focus on respiratory diseases. A questionnaire survey from the rare diseases clinical research network based in Ohio, United States, collects cross-sectional information on the effect of COVID-19 from people with all types of rare diseases (28). A major strength of this study is that recruitment is not happening through health care facilities but any person with a rare disease and access to the internet can participate via a link on the study website. This also ensures that people with mild disease or limited access to health care may participate. However, the study is limited to the United States, the questionnaire is short and collects information only once, and it remains general as it includes people with any kind of rare diseases. Another research project, the ImmunoCOVID19 study, at the University of Southampton, UK, monitors COVID-19 in children with primary or secondary immunosuppression (below 18 years) (29). Recruitment happens through hospitals. Participants complete a short baseline questionnaire and are followed up weekly through a short online questionnaire. This study follows participants longitudinally which makes it fit for studying incidence of COVID-19 and study symptoms and prognosis. Other studies on people with rare respiratory diseases use data from hospital records. The European Cystic Fibrosis Society set up the COVID-CF project in Europe where data from national Cystic Fibrosis (CF) registries are pooled to identify people with CF who have been infected with SARS-CoV-2 (30, 31). By October 04, 144 cases of COVID-19 in patients with CF were documented in 18 countries. A study in Spain investigated incidence of SARS-CoV-2 from March to May 2020 in 2500 CF patients registered in the national registry using information from hospital records (32). However, these studies only identify the most severe cases of SARS-CoV-2 infections that result in hospitalisations. Mild cases are missed, and this type of study biases the results towards more severe infections.

### Strengths and limitations

A key feature and major strength of this study is the participatory approach where people with PCD together with the team of researchers shape the study, recruit participants, and interpret and decide which results should be presented on the study website. The close collaboration between people with PCD and researchers ensures the relevance for participants and makes sure that study material is meaningful for people with PCD. It may also positively influence response rate in the weekly follow-up questionnaires. Another strength is the web-based study design which allows participants to contribute their data to the database in real time. This limits the risk of recall bias as participants only report symptoms from the last seven days, and there is no delay caused by data entry as participants enter data themselves into the database. The PCD support groups provided essential inputs to the study questionnaires and study design. The broad inclusion criteria allowing people of any age from anywhere in the world to participate is another strength of the study. The COVID-PCD study is one of the largest international studies including data reported directly from people with a rare disease. COVID-PCD not only covers infection rates, severity of symptoms, and prognosis of COVID-19 but also studies factors related to the COVID-19 pandemic such as social isolation and physical activity habits. Additionally, special topics can be studied at short notice using extra questionnaires that are sent out once and can be proposed by participants themselves.

One limitation of the study is the convenience sampling method. It is difficult to estimate the response rate without information on how many people with PCD have seen the study information and it is difficult to know if the study participants are representative of the average population of people with PCD.

## Conclusion

The COVID-19 pandemic evolves quickly and gathering data fast on how COVID-19 affects people with rare chronic diseases is essential for informing patients, physicians, and policy makers. First results on recruitment from the COVID-PCD study show a good representation from many regions in the world across all age groups. COVID-PCD is an essential resource to follow people with PCD throughout the COVID-19 pandemic, help empower patients, and in providing a platform for communication between patients, physicians, and researchers.

## Data Availability

Researchers who wish to use data should submit a concept sheet describing the planned analysis and send it to Prof. Claudia Kuehni. The concept sheet will be discussed with the patient advisory group and if agreed, a partial dataset will be prepared by the research team.

## Acknowledgements

We thank all participants and their families and we thank the PCD support groups and physicians who have advertised the study.

